# A multi-center phase III randomized control trial to evaluate effectiveness of the Both EARS (BEARS) virtual reality training package to maximize hearing abilities in children and young people with bilateral cochlear implants: the BEARS protocol

**DOI:** 10.64898/2026.08.12.26360324

**Authors:** Deborah Vickers, Lisabeth Buelt, Liz Arram, Lorenzo Picinali, Marina Salorio-Corbetto, Kashfia Chowdhury, Caroline S Clarke, Nick Freemantle, Dan Jiang, Bhavisha J Parmar, Frances Early, Sandra Driver, Ekaterina Bordea, Teresa Hill, Helen Cullington, Fiona Kukiewicz, Christine Rocca, Padraig T Kitterick, Fleur Corbett, Ruth Nightingale, James Blackstone, Norin Ahmed, Sarah Somerset, Nejra van Zalk, Merle Mahon

## Abstract

**Introduction:** Deafness is the most common human sensory deficit. Cochlear implantation is the primary intervention for severe-to-profound deafness. Currently, over 7000 people have bilateral cochlear implants (CIs) in the United Kingdom (UK), most of whom are children. Patient feedback suggests that for children with bilateral CIs, everyday communication is challenging and tiring, with extra effort required to integrate information from two ears, especially in noise, and that current rehabilitation techniques are not engaging, or appropriate to their lifestyles. To address these issues, researchers developed the Both EARS (BEARS) training package comprised of three virtual reality games to improve sound localization and listening in noise.

**Objectives:** This protocol describes the design and methodology of a multi-center phase III randomized controlled trial (RCT) to evaluate whether use of the BEARS training package alongside usual care compared to only receiving usual care improves speech-in-noise perception, hearing experiences, vocabulary and quality of life and reduces listening effort in children and young people (aged 8 -16 years (inclusive) with bilateral CIs.

**Methods:** This RCT is currently underway in 16 clinical CI departments in National Health Service or university hospitals across the UK. The intervention involves 3 months of spatial-listening training delivered via the BEARS training package in addition to any routine rehabilitation. The control is ‘usual care’ (routine rehabilitation clinical care pathway). The primary outcome is the difference between the intervention groups in speech-in-noise perception score at 3 months derived from the spatial speech in noise (SSiN-VA) test. Recruitment closes at the end of the day on 31^st^ July 2026, and end of data collection is 31^st^ October 2026. Data analyses will be reported by 31^st^ March 2026.

**Significance:** This is the largest known trial of children and young people with bilateral CIs. It will generate high-quality evidence on speech-in-noise outcomes and inform training interventions to improve spatial listening.

**Trial registration:** ClinicalTrials.gov registration: NCT05808543; UK’s clinical study registry (ISRCTN92454702)

## Introduction

Cochlear Implants (CI) are one of the most successful medical devices available. They are surgically implanted and contain an electrode array inserted into the cochlea that bypasses the damaged cochlear hair cells and stimulates the auditory nerve directly. CIs are the main hearing device intervention for people with severe/profound sensorineural hearing loss. In the UK there are currently 18,300 adults and 6,600 children using at least one CI and there are over one million worldwide [1]. With respect to deaf children, the National Institute for Health and Care Excellence (NICE) has recommended bilateral CIs since 2019 [2], because the benefits of hearing through two ears greatly outweighs the use of only one. There are currently 5,500 children with bilateral CIs in the UK with approximately 400 new children bilaterally implanted annually [1].

The primary intention of CIs is to provide access to speech (to develop spoken communication and acquire language skills for children) for the deaf listener. Speech communication is crucial for cognitive, social and language development and often takes place in complex noisy environments. For listeners with normal hearing, subtle differences in timing and level of sounds reaching each ear provide directional and localization cues for ‘spatial hearing’ [3–6] which contribute to the separation of speech from noise and the ability to attend to a particular speaker [7–9].

Children and teenagers spend substantial time in noisy environments, impacting their speech and language perception, educational attainment, and social well-being. Hearing loss further adds to these challenges and increases the risk of emotional and behavioral difficulties that can influence social relationships [10,11]. Although language development [12–14], sound localization and tracking of moving sound sources [15–20], speech-in-noise perception [14,15,21] and listening effort [22] are better for people with bilateral CIs than for those with a unilateral implant [23,24], these skills remain far below those with normal hearing. People with bilateral CIs often struggle to understand speech in noise even with visual cues [25].

Our ‘Living with CIs’ Patient and Public Involvement and Engagement (PPIE) group reported that everyday communication was challenging and tiring due to the extra effort required, especially in noise [26]. Bilateral CI users reported how it can be hard to balance sounds from two implants and they described sounds from the second implant as ‘annoying’, ‘distracting’ and ‘lop-sided’, leading to its rejection.

The lack of protocols for fitting bilateral CIs, ecologically valid outcome measures, and resources for spatial-hearing training contribute to these listening difficulties. The current ‘usual care’ rehabilitation assessments and techniques completed with bilateral CI patients are reportedly not engaging, do not reflect real-world hearing difficulties experienced by users and do not feature specific training to make the most of having two implants [26,27]. However, a large body of research demonstrates that sound localization can improve with training, underpinned by plasticity-driven changes in the auditory pathways for children and adults [20, 28–45]. Additionally, research has found that computer-based speech training can improve sentence recognition [46] and that simple activities in which users interact with sounds through movements and vision can result in significant hearing improvements [47]. Pediatric CI users have also reported that computer-based rehabilitation assessments would better fit their lifestyles and support their learning [26].

To help address the everyday hearing challenges faced by children and young people with bilateral CIs and to reduce current gaps in rehabilitative support for this population, we developed the Both Ears (BEARS) training package of virtual-reality (VR) games [27] to train spatial hearing. A participatory design approach was used to develop the training package [26], improve its relevance to the target population and enhance the likelihood of sustained use.

The BEARS training package comprises three games addressing different hearing functions: speech-in-noise perception, music listening and sound-source localization. Each game is based on an audiovisual task performed through a VR interface. The BEARS training package uses a head-mounted display, such as the Meta Quest 2, or an iPad for those with balance issues or who have a head circumference of less than 50 centimeters. Audio is delivered, in both cases, using circumaural headphones (AKG K240 Studio). Different audio playback options were widely explored during the participatory process [26] including using wireless audio transmission. Due to the high variability across CI manufacturers, and potential latency and quality issues with streaming audio via Bluetooth, the choice of the circumaural headphones was ultimately taken, and the brand and model were chosen together with the various stakeholders.

By providing training using videogames that are engaging, relevant and appealing to young people, it is expected that positive brain changes can occur to maximize spatial-hearing skills. This has the potential to improve auditory performance in real-life scenarios, which would have positive repercussions in social, emotional, and academic aspects of life. To date, no training program has been implemented and evaluated for young people with bilateral CIs. This manuscript describes the design and methods of a large RCT that aims to evaluate the effectiveness of the BEARS VR training package in improving hearing abilities and listening skills in children and teenagers with bilateral CIs.

An independent, parallel mixed-methods process evaluation sub-study, that collects data from participants, caregivers, and clinicians, will run alongside the trial. Designed to support interpretation of the trial results, the process evaluation will inform optimization of the intervention and identify strategies for successful implementation [48], guided by Normalisation Process Theory [49,50]. The process evaluation will assess the quality of trial implementation including fidelity, dose and reach, the participant experience of using BEARS and contextual factors associated with variations in outcome, as well as gathering clinician experiences of implementing the trial. The process evaluation will make recommendations for intervention delivery and will be reported separately.

### Objectives

The primary objective of the BEARS trial is to determine whether spatial-listening training for three months delivered via the BEARS training package in addition to usual care compared to usual care alone, improves spatial speech-in-noise perception when assessed at three months (accounting for baseline).

The secondary objectives investigate the differences between the intervention and control trial arms and explore whether the BEARS training package (used for three months):

1. Improves spatial speech-in-noise perception when assessed at twelve months (accounting for baseline)
2. Improves localization abilities when assessed at three and twelve months (accounting for baseline)
3. Reduces listening effort when assessed at three and twelve months (accounting for baseline)
4. Increases vocabulary age when assessed at twelve months (accounting for baseline)
5. Improves quality of life when assessed at three and twelve months (accounting for baseline)
6. Is cost effective when assessed at three and twelve months (accounting for baseline)
7. Improves perceived benefits of everyday listening when explored at three months compared to baseline
8. Improves music listening experiences when explored at three months compared to baseline.

## Methods

### Patient and public involvement and engagement (PPIE)

PPIE work has shaped the trial from the development of the initial research question, the design of the intervention, the planning of the trial design and the communication of outcomes [26]. This was supported by the Cambridge Biomedical Research Centre PPIE Group, and all activity follows UK standards for Public Involvement [51].

Multiple PPIE groups made up of deaf children and young people, clinicians and therapists, teachers and families have informed the design of this work. These groups are inclusive and contain a diverse range of people from different ethnic and socio-economic backgrounds. Furthermore, during the trial, PPIE groups have supported the trial team with study documentation (consent/assent forms, information sheets) to ensure that they are accessible to diverse participant groups. They have also collaborated on the design of data collection forms for resource use, provided recruitment ideas to enhance enrollment, and advised on website design and dissemination as well as the design and delivery of the qualitative work and process evaluation.

### Trial Design

The effectiveness of the BEARS training package is being evaluated in a multi-center, randomized controlled, phase III superiority trial. Participants will be randomized in a 1:1 ratio. BEARS is a non-CTIMP (Clinical Trial of Investigational Medicinal Product) study. The trial is being conducted from all 16 hospital-based pediatric CI centers in the UK. The BEARS intervention is home-based training.

This protocol was developed using the Standard Protocol Items: Recommendations for Trial Interventions (SPIRIT) checklist [S1 File] and participant timeline [Figure 1] and reflects version 7 of the study protocol approved by the ethics committee [S2 File].

**Figure 1.**
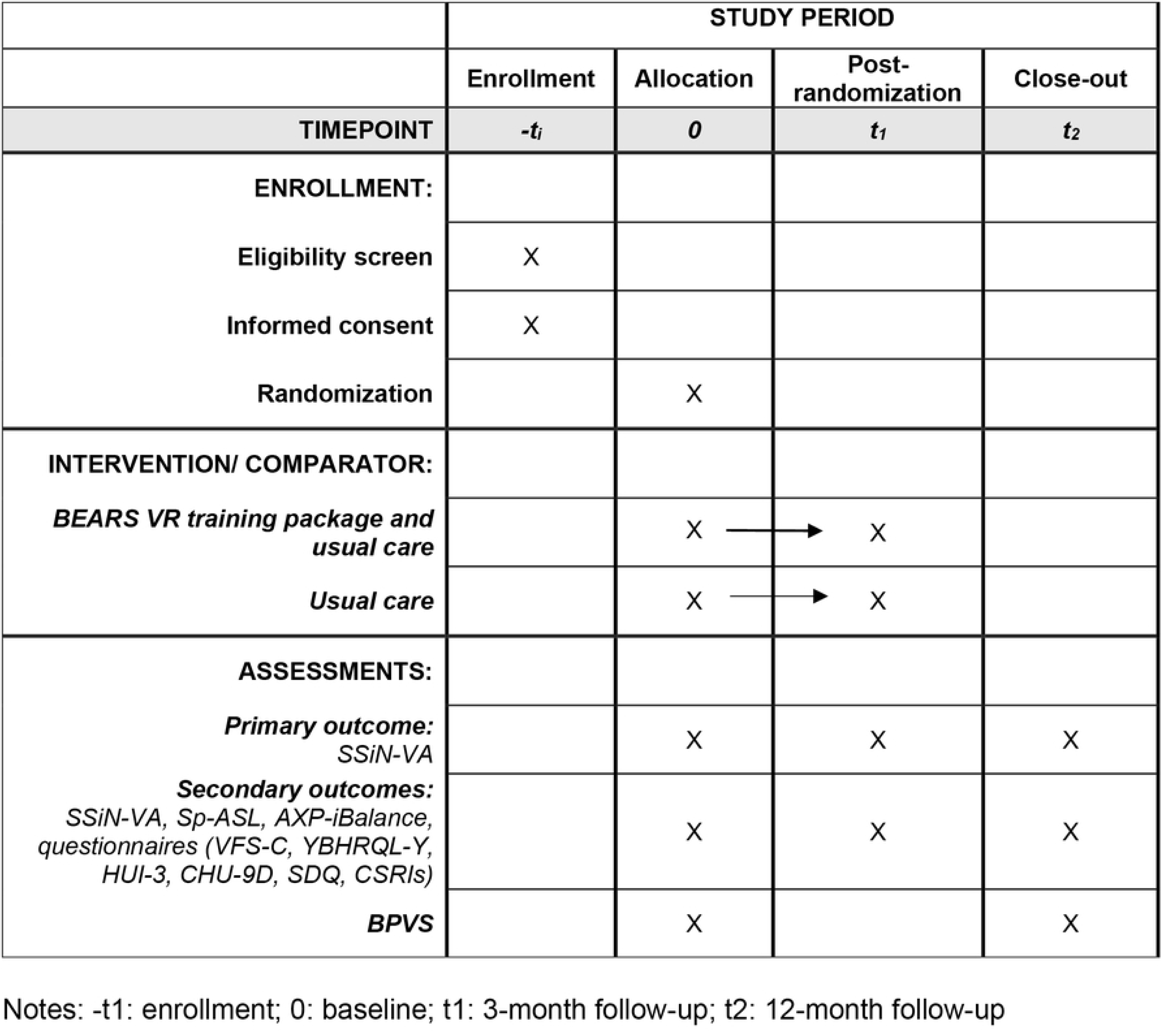
Participant timeline.

### Status of study

The date of first patient enrolment was 4 August 2023 with the final enrolment by end of the day on 31 July 2026 and last-patient-visit 31 October 2026, with the project ending on 31 March 2027 and first data analyses will be reported by this date.

### Site eligibility

All pediatric CI centers in the UK are potentially eligible and have been approached prior to the trial opening to recruitment. They have been assessed to determine whether they could feasibly run the trial, have the capacity to do so (staff and space), and to establish the numbers of potential participants for recruitment. The Principal Investigator (PI) at each site does not need to be a medical member of staff.

### Participants

Participants will be considered eligible for enrolment in this trial if they fulfill all the inclusion criteria and none of the exclusion criteria as defined in Table 1.

**Table 1.** Inclusion and Exclusion Criteria for the BEARS study.

| Inclusion criteria |  |
| --- | --- |
| **To be eligible, a participant must:** |  |
| 1. | have simultaneous or sequential bilateral CIs, using both CI processors for a minimum of 6-hours per day (measured over a month), who either has:<br>a. Congenital severe/profound bilateral sensorineural hearing loss and have received at least one implant ≤ 36 months of age.<br>b. Progressive or acquired severe/profound bilateral sensorineural hearing loss (no age at implant restrictions for these patients). |
| 2. | have stable CI programs (defined as no longer using progressive programs to work through). |
| 3. | have had at least two usual care checks/clinical appointments with stable aided levels (± 10 dB across 500Hz-4kHz) and no progressive maps to still work through, if they have had re-implantation of internal implant devices |
| 4. | be aged in the range 8-16 years, inclusive. |
| Exclusion criteria |  |
| **Participants are excluded if they:** |  |
| 1. | do not (or caregivers do not) speak/understand English sufficiently to undertake assessments. |
| 2. | have an intellectual disability at a level that would prevent their ability to understand the trial the intervention or assessment questions. |
| 3. | have a comorbid condition impacting ability to participate in the intervention and/or outcome assessment. |
| 4. | have an audiological profile impacting ability to participate in the intervention and/or outcome assessments. |
| 5. | Are actively participating in other trials that may affect hearing outcomes or impact their ability to participate in the intervention. |
| 6. | are currently or anticipated to receive treatment and/or intervention that may affect hearing outcomes or adapt implant settings/programming. |
| 7. | refuse to consent to trial activities/protocol. |
| 8. | Are awaiting reimplantation following device failure or infection. |
| 9. | Are a non-user of one or both implant processors (i.e., must use both processors for a minimum of 6 hours per day over a month). |
| 10. | are a full-time boarder at a boarding school |
| 11. | have unresolvable issues found in device checks that render one of the implants unusable. |
| 12. | are pregnant.*<br>*\*If a participant becomes pregnant during the trial they can remain in the trial because the BEARS training program does not pose a risk to the development of the fetus.* |
| 13. | have a diagnosis of epilepsy or history of seizures of any kind. |

### Interventions

#### Intervention arm: BEARS training package

The BEARS training package is a suite of three videogames designed specifically for young people with bilateral CIs. The hardware is either:

- A head-mounted VR display device (Meta Quest 2) with AKG K240 Studio headphones; or
- An iPad with AKG K240 Studio headphones, for those participants with a smaller head circumference (<50 cm) or with reported balance conditions.

The BEARS training package comprises three games addressing different hearing functions: speech-in-noise perception, music listening and sound-source localisation. Each game is based on an audiovisual task, and participants are automatically guided through on-screen visual prompts to support the gameplay with feedback given on their performance and progress through levels of increasing difficulty.

The three games are:

1. **Target training game:** Initially developed to train normal hearing listeners to localize sounds [40], it has subsequently been adapted for CI users. Through a series of increasingly difficult levels, the player is trained to localize sounds relying initially on audiovisual cues. The visual signal disappears gradually as the levels increase, transforming the task into a purely auditory challenge. Steadman et al. have shown how virtual auditory displays can be used for creating training environments where users learn to localize sounds using modified localization cues [45]. The use of audiovisual stimuli helps with task familiarization in the lower levels, and the gamification approach contributes to improving engagement and attainment.
2. **Speech-in-Noise training game:** Follows many of the same principles as the localization game. It is based on the principle that computer-based training of word perception in noise can improve speech perception [52]. Players will be introduced to a virtual scenario simulating everyday situations, e.g., café. A series of increasingly difficult speech recognition tasks will be presented, challenging the players to interact with the environment (e.g., rotate their head), localize the speaker and identify the words, using a range of background noises and reverberant acoustic conditions.
3. **Spatial music game:** This is based on a ‘Musiclarity’ application created within the 3D Tune-In project [53]. The game aims to enhance perception and localization of musical instruments and lyrics in a range of immersive and interactive soundscapes.

The BEARS training package design allows for the training to be self-administered, played anywhere and at any time. There is no upper limit to the frequency of use of the BEARS training package. During each session of use (minimum 2x 30-minute sessions once a week), all three games need to be played. The device steers participants through the three games and participants receive an instructional guide that describes how to use the Meta Quest 2 headset [54].

The participants will also be provided with a training diary to document any issues they have with the BEARS training package hardware or software, and to document when the games had been played, helping to ensure compliance with the minimum training. Use of the BEARS training package requires all participants randomized into the BEARS intervention arm to switch to an extra program added onto their CI sound processors. This program removes microphone preprocessing modifications (e.g., directional microphone, wind noise reduction). Once they have finished using the games, they then return to their everyday program.

#### Control intervention: Usual care

Usual care describes the routine rehabilitation received by participants via their CI center. During a review appointment the clinician will check that all external and internal equipment is working and check CI usage logs. Patients can attend their CI center at any time to discuss any equipment issues. This will enable their CIs to keep working optimally according to the British Cochlear Implant Group (BCIG) quality standards (QS) [55]. There is no limit to the level of contact between the patient or their family/legal representative and the implant center, in line with BCIG QS. Any contacts will be recorded in the patient’s medical notes and appropriate sections of the trial’s case report forms (CRFs).

### Concomitant care

Treatment for any other conditions will be given as per the usual care pathways. Eligible BEARS participants will not be enrolled if they are currently receiving, or are anticipated to receive, treatment and/or an intervention that could modify their CI settings or affect hearing outcomes.

### Criteria for modifying allocated intervention

If a participant in the BEARS intervention arm is unable to use the head-mounted display device for any reason, they will be provided with an iPad alternative. Participants in the BEARS intervention arm who report serious adverse events which are deemed related to the trial intervention may continue with the intervention, if consenting to, at the discretion of the site PI. Where serious adverse events continue to be experienced, the participant, their parent/legal representative, and the investigator at the trial site should decide on whether to stop the trial intervention. If the trial intervention is stopped, the participant should continue to be followed for the remainder of the trial duration unless they explicitly withdraw their consent for follow-up. If a participant on the BEARS intervention experiences an interruption or discontinuation associated with either or both CIs, requiring implantation or device setting alteration, they will discontinue the trial intervention and remain in trial for follow-up (as per intention-to-treat analysis principles).

A subset of participants will take part in qualitative interviews. An independent process evaluation runs alongside the trial to gather data on fidelity, compliance, participant experience and contextual factors associated with variations in outcome.

### Consent and screening procedures

Potential participants will be identified through pre-screening measures completed at the implant center where they are receiving usual care. The clinical team will review potential participants’ clinical notes, evaluating their implant history and demography, and document in a pseudonymised pre-screening log.

Following pre-screening, the clinical implant center research team will contact eligible trial participants and provide appropriate parent and patient information sheets (PIS) that describe the trial details, procedures, and risks in simplified form (these sheets are available on the BEARS website). The PIS will be provided to potential participants and their parents/legal representatives in advance of the online consenting visit (Visit 1). They must be given sufficient time to read and understand the PIS (minimum of 24 hours) and be given the opportunity to ask questions and discuss the aims, methods, benefits, and potential hazards of the trial, as well as the optional qualitative interviews as part of the sub-studies (listed below) before providing electronic informed consent and/or assent. Following Health Research Authority (HRA) guidance, patients under 16 years of age will provide assent alongside the parent/legal representative consent and participants who are 16 years old, will provide informed consent alongside their parent/legal representative. The person taking consent/assent will need to have had GCP training [56], be suitably qualified and experienced, and have been delegated this duty by the PI on the delegation log.

To promote recruitment across sites, the Trial Manager will be in contact with the site teams on a regular basis to provide support for any recruitment questions. Additionally, the Trial Manager will host online ‘Tea break’ sessions every two weeks, providing site teams with a forum to receive trial updates, share successful recruitment strategies, and pose trial-related questions. The Chief Investigators will arrange motivational site visits with the Trial Manager and other members of the team, and the Programme Steering Committee Chair will contact sites to encourage involvement.

To promote retention in either study arm, participants will be given gift vouchers upon completing each follow-up appointment. Participants will receive £10 upon completion of the baseline, and £20 upon completion of the 3 and 12-month visits. Additionally, travel expenses for the 3-month visit will be compensated. If the participant is randomized to the usual care arm, they will have the opportunity to sample the BEARS training package once they have completed all their follow-up appointments.

### Randomization

Randomization will be performed by the PI or delegated member of the trial team at local sites using Sealed Envelope, an independent online randomization service that minimizes allocation bias within the trial [57]. This has been validated by the trial statistician. Following confirmation of eligibility and consent, each participant will be randomized using their screening identification number that was allocated sequentially at screening.

### Type of randomization

Participants will be randomized 1:1 using a concealed fixed minimization process according to the minimization factors listed below. This will ensure that balance across the two study arms will be achieved. The minimization algorithm will incorporate a random element to maximize balance in the stratifying variables between the randomized groups.

Minimization factors will include: 1) sex at birth (male; female); 2) school age group (primary 8-11; secondary 12-16); 3) school setting (mainstream day school; weekday specialist boarding); 4) onset of deafness (congenital; progressive/acquired); 5) device allocation (VR headset; iPad).

Sealed Envelope software ensures that site clinicians are unaware of the next assignment in the random sequence. For the sites, clinicians’ allocation concealment is not possible during the intervention period because the equipment/training aspects of the BEARS intervention need to be provided to the participant and their family at the end of their baseline visit (Visit 2b).

Responsibility for enrolling participants and randomizing them to study arms lies with the PI and other trained individuals (e.g. clinicians/research nurses) named on the site delegation log.

Once a participant has been allocated to a study arm and following baseline data collection, the participant/family will be informed of the intervention allocation by a delegated site team member.

### Blinding

This trial is unblinded. It is not possible to blind patients, clinicians and researchers to the intervention allocation following randomization. However, the Chief Investigators, the central team running the trial and the main statistics and health economics team will remain blinded to participant allocation until recruitment has ceased.

### Sample Size

The sample size is based on the ability to detect the clinically important difference between treatment groups of 2.25 percentage points change in speech-in-noise-perception score on the SSiN-VA test [58]. This has 80% power at the 5% level of significance (two-sided). The standard deviation (SD) of change from baseline is estimated to be 6.05 percentage points. Mitigating loss to follow up or other challenges, a 15% allowance requires a sample size of 136 per group, totaling 272 participants (the original version of the protocol proposed recruitment of 384 participants but this was adjusted to a lower power (80% from 90%) and as participant retention was higher/better than predicted the attrition rate was adjusted (to 15% from 20%). These changes are detailed in the Summary of Protocol Amendments document [S3 File].

### Sub-studies

At Visit 1, all participants will be offered the opportunity to take part in interviews. Participants who provide consent may be contacted for either a qualitative sub-study interview or a qualitative process evaluation interview. For the longitudinal qualitative sub-study, a minimum of 20 participants from each study arm will be interviewed online via Zoom at baseline and again after three months about their everyday hearing experiences before and after taking part in the trial. In addition, all participants in both arms of the trial are asked to respond to open-ended survey questions about their experiences of hearing in daily life, at successive timepoints throughout the study (baseline, 3 and 12 months).

### Process Evaluation

At Visit 2b, all BEARS intervention arm participants will complete a questionnaire about their confidence in using, motivation to use, and expectations of, the BEARS intervention. At Visits 3a and 4a, all BEARS intervention arm participants will complete further questionnaires about their experience of using BEARS and the context in which BEARS was used. Up to 20 participants from the BEARS study arm will be purposively sampled for representation across age and sex and interviewed on Zoom at three months, about their experiences of using the BEARS training package.

At Visit 2b, Parents/legal representatives will complete questionnaires about prior technological access and engagement and the Strengths and Difficulties questionnaire [59]. Parents/legal representatives will answer questions about their experience of the trial at Visits 3a and 4a.

Clinicians named on the delegation log at each clinic site will be invited to complete the NoMAD questionnaire [60] at the first participant’s Visit 2a and Visit 4b. Clinicians’ experiences of trial delivery and factors relevant to implementation will be collected from case study sites identified for maximum variation of: healthcare system, NHS and non-NHS providers, number of patients and clinicians, location, prior research experience, trial delivery by local clinicians or research team.

Online focus groups and/or interviews will be conducted with up to eight clinicians from each case study site at the site’s first Visit 4b.

### Outcomes

#### Primary Outcome

The primary outcome for the BEARS trial is the difference between study arms in speech-in-noise perception score (% correct overall task) at 3 months, accounting for the participant’s baseline score. This is derived from the SSiN-VA test [58], a dual-task assessment that simultaneously assesses word identification and relative localization and can provide information about spatial release from masking. The SSiN-VA is based on a test initially developed by Bizley et al. [61] and has been adapted for virtual implementation [58], eliminating the need to use a multi-speaker array to assess spatial listening.

#### Secondary Outcomes

- **Additional SSiN-VA test outcomes:**

○ Speech-in-noise perception score (% correct of the overall task) at twelve months accounting for the baseline score.
○ Relative localisation score (% correct) at three months and at twelve months accounting for the baseline score
○ Average reaction time (measure of listening effort) for word identification selections at three months and at twelve months accounting for the baseline score.
○ Average reaction time (measure of listening effort) for location shift selection at three months, and at twelve months accounting for the baseline score.
○ Spatial index for word identification at three months and at twelve months accounting for the baseline score.
○ Spatial index for relative localization at three months and at twelve months accounting for the baseline score.
- **Spatial Adaptive Sentence List (Sp-ASL) test outcomes.** The Sp-ASL, is an adaptative speech test and a standard outcome measure [62] administered in accordance with the Bamford-Kowal-Bench Speech-in-Noise (BKB-SIN) task protocol [63] This test will capture:

○ Speech reception threshold at three months and at twelve months, accounting for baseline (for better ear, worse ear, and average of both).
○ Spatial release from masking score at three months and at twelve months, accounting for baseline (for better ear, worse ear, and average of both).
- **British Picture Vocabulary Scale (BPVS) 3^rd^ edition test outcome.** The BPVS is a validated vocabulary assessment that can assess a child’s receptive (hearing) vocabulary [64]. This trial will assess the difference between arms in vocabulary age at twelve months, accounting for baseline vocabulary age.
- **Vanderbilt Fatigue Scale: Child self-report version (VFS-C) questionnaire outcome.** The VFS-C is a unidimensional scale designed to assess listening-related fatigue in children ages 6-17 years [65]. This trial will assess the difference between arms in listening-related fatigue score at three and 12 months, accounting for baseline.
- **Health economic and quality of life outcomes.** The economic evaluation will calculate incremental cost per quality-adjusted life-year (QALY) gained by offering BEARS and usual care, compared to usual care, from an NHS, Personal Social Services (PSS) and Local Education Provider perspective over the twelve months of the trial. Mean pathway cost per participant in each arm will be calculated by applying standard unit costs to resource use information captured and adjusting for baseline values (i.e., Client Service Receipt Inventory questionnaires (CSRI), plus resources and costs associated with the intervention as applicable). Mean QALYs over 12 months per participant in each arm will be calculated, adjusting for baseline utility values, from responses to the self-completed York Binaural Hearing Related Quality of Life questionnaire for young people (YBHRQL-Y), proxy-completed Health Utilities Index (HUI-3) and the self-completed Child Health Utility 9 Dimensions (CHU-9D). The YBHRQL-Y is a validated self-complete questionnaire [66] adapted from the YBHRQL [67], and is a hearing-specific, preference-based-measure of quality-of-life in children. The HUI-3 is a system used to measure generic health-related quality of life and has demonstrated significant sensitivity to hearing loss [68]. The CHU-9D is a validated, pediatric generic preference-based measure of health-related quality of life [69].
- **Process evaluation outcomes.** BEARS participant’s intervention use will be automatically logged within the BEARS games for calculating dose. Reach will be measured with participant implant data and sociodemographic information. Participant confidence, motivation and user experience, technological engagement and home environment will be measured through questionnaires to explore the effect of context on the primary outcome and BEARS datalogs. The SDQ [59] completed by parents/legal representatives measures a child’s specific strengths and difficulties. The NoMAD [60] measures clinician’s views towards BEARS and their willingness to incorporate it into routine practice. The participant experience of using BEARS and the clinician experience of delivering BEARS will be captured in interviews and focus groups.
- Qualitative outcomes relating to how experiences of everyday/real-life hearing and listening (including music) change over time and might be different between intervention groups.

#### Exploratory outcomes

- **Age effects** [For all outcomes across the 12-month trial involvement]. No further data will be collected for the age effect analyses but the data regarding the trajectory of the children’s speech-in-noise scores relative to normative age range for the speech measures will be assessed to determine if there is a difference in trajectory over time between the BEARS intervention and usual care groups. These same profiles will be explored for all the outcome measures in the trial to determine how the child’s age (chronological, developmental age based on vocabulary level and hearing age based on numbers of years hearing with an implant)
- **Retention of training effects**. No further data will be collected for the retention effect analyses. It is considered best practice in training interventions to evaluate if the training effects remain after the intervention period has finished. For this analysis fixed effects of timepoint (baseline, 3, and 12 months) and group (BEARS intervention, usual care) and random effect of participant will be used to understand the speech-in-noise outcomes.
- **Impact of degree of balance between ears.** For this analysis, the AXP-iBalance application will be used to indicate the degree of symmetry across the two ears. The balance measurements at baseline, three months and 12 months, will be used as an outcome to determine if there have been any changes in the balance between the ears over time.

### Safety considerations and strategies for reducing harm

Use of the BEARS training package is low-risk. Potential risks associated with the use of the Meta Quest 2 VR head-mounted display are documented in the Meta Quest 2 for business safety guide [70]. By having eligibility criteria for potential participants and instructions on how and when to use the equipment, these risks are greatly reduced. If symptoms of pain, discomfort, prolonged dizziness, imbalance or nausea are reported, the participant can complete the intervention using an iPad.

The pediatric participant population for the BEARS trial is a higher risk group because of the potential association between hearing loss and balance issues and the higher chance of multi-comorbidities. However, due to the current clinical pathway, participants approached to be recruited onto the trial will be well known to the clinical teams who will be aware of any balance issues or comorbidities that could affect engagement in the trial or whether the use of an iPad instead of a VR headset would be appropriate. Assessments will be completed by appropriately trained staff at the clinical sites.

It is considered that the potential benefits of the BEARS training package outweigh the aforementioned potential risks.

The principles of GCP [71] require that both Investigators and Sponsors follow specific procedures when notifying and reporting adverse events or reactions. Adverse events in this context include exacerbation (i.e., increase in the frequency or intensity) of a pre-existing illness, episodic event or symptom (initially recorded at the baseline visit), that is detected after intervention, or occurrence of a new illness, episodic event or symptom, that is detected after intervention.

Adverse events do not include medical or surgical procedures, pre-existing disease or a condition present before treatment that does not worsen, hospitalization where no untoward or unintended response has occurred, or overdose of medication without signs or symptoms.

From the time of consent, all pregnancies and suspected pregnancies that occur in participants must be reported immediately to the clinical trials unit upon the site becoming aware. Pregnant participants must be given a copy of the trial pregnancy monitoring information sheet and asked to complete a pregnancy monitoring informed consent form for follow-up in pregnancy [S4 File]. The process should follow GCP, and the processes used to obtain consent for the clinical trial.

If a participant who is randomized to the BEARS intervention arm becomes pregnant during the first three months of trial, they must immediately discontinue use of the Meta Quest 2 and will be provided instead with an iPad to complete the BEARS training period. Any complications of pregnancy including miscarriage, congenital abnormality or birth defect resulting from the pregnancy must be reported as a Serious Adverse Event (SAE).

### Data collection, management, and analysis

#### Data collection methods

Trial data will be stored in a database created specifically for the BEARS trial hosted by OpenClinica, USA [72], which is validated for 21 Code of Federal Regulations, Part 11, GCP, Health Insurance Portability and Accountability Act, General Data Protection Regulation (GDPR), and Annex 11. The data is stored on secure, GDPR-compliant, cloud-based servers held within UK and European Union [73]. Trial data will be collected at the timepoints indicated in the Participant Timeline (Figure 2). For this trial, the majority of datapoints will be recorded directly into electronic CRFs in the OpenClinica database. Data collection will only occur by delegated members of the trial team across participating sites.

**Figure 2.**
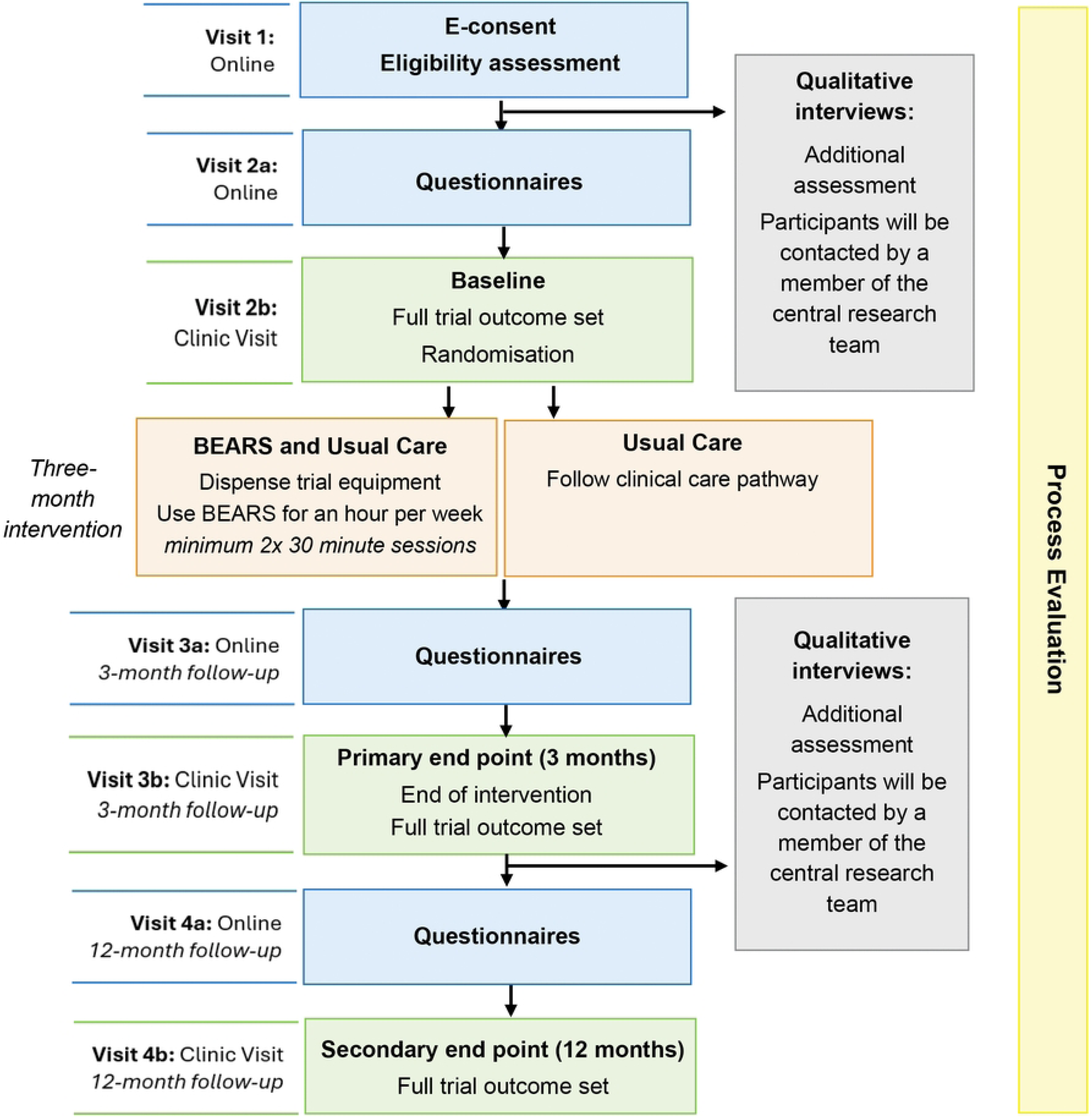
Participant Flowchart.

During clinic visits, trained members of the trial team will administer the three iPad-based BEARS applications - the SSiN-VA, Sp-ASL and AXP-iBalance - and send the results to the research team. The ‘BEARS Clinician Guide’ document provides instructions on how to carry out these assessments, and how to extract data to send to the research team [74]. Additionally, a delegated member of the trial team will conduct the BPVS at timepoints identified in the Participant Timeline (Figure 2). Data are recorded in the participant’s medical record and entered in the electronic CRFs in OpenClinica.

The BEARS device will automatically log progression and frequency of use, and the player’s performance scores for later analysis. No sensitive or personal data is saved onto the device. Once the 3-month intervention has been completed and the device has been returned to the clinic, all data will be downloaded from the device and then deleted. For participants who withdraw from the trial, no further information will be collected from the date of withdrawal.

NoMAD Process Evaluation data will be collected from clinicians electronically via the Qualtrics GDPR compliant platform.

#### Data quality and control

The OpenClinica database software helps maintain data quality, including maintaining an audit trail, allowing custom validations on data, allowing users to raise data query requests, and search facilities to identify validation failure and missing data. Data collection, data entry, queries raised by a member of the BEARS trial team and database lock(s) will be conducted in line with the CCTU SOPs and BEARS Data Management Plan. All data will be handled in accordance with the Data Protection Act 2018, the UK General Data Protection Regulation (UK GDPR) and subsequent updates and amendments.

#### Data security

The randomization service is hosted by Sealed Envelope LTD [75]. The data is stored on a secure, GDPR-compliant, cloud-based servers held within the EU. These databases are protected by multi-layer firewalls with full data encryption at rest and in transit. The identification, screening, and enrollment logs, linking personally identifiable information to the participant identification number (PIN), will be held locally by the trial site. This will either be held in written form in a locked filing cabinet or electronically in password protected form on hospital computers.

Data that is external to the trial database, such as training and compliance records for those in the BEARS study arm, the raw SSiN-VA, Sp-ASL, AXP-iBalance application, data logging data, process evaluation and qualitative data will be saved in password protected secure form on institutional computer systems.

#### Data storage

Central review of the SSiN-VA, Sp-ASL and AXP-iBalance assessments will be undertaken by the central trial team at Cambridge University on behalf of the sites. These datasets will be stored on the University of Cambridge OneDrive. Qualitative datasets will be stored on UCL OneDrive.

After trial completion, the database will be retained for ongoing analyses. The CCTU will retain a copy of the database, which will be password protected and only accessible to members of the BEARS trial team at CCTU, delegated site staff and external regulators if requested. Database users will only be granted permission to use the database functionality appropriate to their role in the clinical trial. Once all primary, secondary and health economic and exploratory analyses have been completed, the trial data will be archived. Once the trial data has been archived, the trial database will be decommissioned and will no longer be available. Any subsequent or further analysis will be performed using the archived data. This will be within one year of the end of the trial.

The investigators agree to archive and/or arrange for secure storage of BEARS trial materials and records/essential documents for a minimum of 5 years after the close of the trial unless otherwise advised by the CCTU or Sponsor.

#### Confidentiality

We will follow the principles of the UK Data Protection Act 2018 and subsequent amendments. Participants’ data will be collected and kept securely. Confidentiality of participants’ personal data is ensured by not collecting participant names and other personally identifiable information on CRFs and receiving only pseudonymized data. Copies of participants’ trial data will be kept at the participating site in a secure location with restricted access.

To ensure confidentiality for the qualitative data, any identifying information will be removed from the transcriptions. When publishing results, care will be taken not to report information that will enable participants to be identified, for example in relation to rare conditions or geographical locations.

#### Data Sharing

Datasets relating to the different outcomes of this trial will be made available at the time of publication. The data will be anonymized, published with participant level results but no information about the clinic where they are seen.

### Statistical and health economics analysis plan

Primary, secondary and exploratory analyses will be conducted by the principles of intention-to-treat. We will use a linear mixed model to compare the speech-in-noise perception scores between the two arms. The baseline and three-month measures will be included as outcome variables. The fixed-effect of the model will include an indicator variable to identify pre-and post-intervention measures and the minimization factors and loudness imbalance between ears, as adjusting variables. A random intercept term will be included to account for clustering within participants, and an additional random intercept will be included to account for clustering by site.

We will fit analogous models with appropriate link functions and error terms for the analysis of the secondary outcomes, speech-in-noise perception at 12 months and BPVS, and VFS-C at three and 12 months.

Quality of life scores (also called “utility scores”) for each arm will be calculated using the newly determined preference-based value set for the YBHRQL-Y (i.e. as determined in work package 5 of the overall programme grant), and the existing value sets for the HUI-3 and CHU-9D. They will be compared across the two arms, adjusting for baseline values.

All randomized participants in each arm will be included in analyses, including available data from participants who have withdrawn.

#### Population Analysis and Missing Data

Analysis will be conducted on all patients randomized to either the BEARS or usual care study arms. Primary outcome analysis will follow the intention-to-treat principle where all randomized participants are analyzed in their allocated group, whether or not they receive their randomized intervention plan. We will conduct sensitivity analyses to examine the potential influence of missing data under different pattern assumptions using joint models.

#### Subgroup analyses

Using interaction terms, we will explore whether a differential treatment effect is observed according to the following baseline factors: sex (male vs female), school age group (primary vs secondary), school setting (boarding during the week vs mainstream), onset of severe/profound hearing loss (congenital or progressive/acquired), whether participants are in criteria for iPads or head-mounted displays, vocabulary delay (no delay/moderate delay vs severe delay) and loudness imbalance between ears.

#### Adjusted analyses

Where appropriate, analyses will additionally adjust for other baseline factors. Further details of these analyses can be found in the statistical and health economics analysis plan [S5 File].

### QoL instrument measurement properties

We will examine associations between the YBHRQL-Y, the VFS-C and other measures of speech in noise perception and listening effort at all measurement timepoints to further assess the construct validity of the YBHRQL-Y in measuring the intended quality of life construct. We will similarly explore differences in the sensitivity to change of the YHBRQL-Y over time compared to the HUI-3 and the CHU-9D.

### Health Economic Analysis Plan

The health economic analysis will be a within-trial cost-utility analysis as outlined above. The statistical analysis plan and the health economic analysis plan are complementary.

### Qualitative data analysis plan

All qualitative data will be imported into NVivo software to facilitate data handling, organization and coding. Data will be analyzed thematically using Framework analysis [76]. All members of the sub-study research team will be involved in reading and coding transcripts. Coding will be both deductive (driven by the research questions, such as understanding speech in noise, localizing sound sources and listening to music) and inductive to ensure emergent data-driven themes are also identified. Additional information about the management of qualitative data can be found in the Qualitative Data Management Plan [77].

All themes will be developed and revised in an iterative manner as patterns within the data become more apparent. Emerging themes will be discussed in regular meetings with the qualitative research team. The analysis will be complemented by discussions with our PPIE groups.

There will be a final mixed methods analysis to integrate the quantitative and qualitative trial outcomes.

### Process Evaluation Analysis Plan

Descriptive statistics will be reported for fidelity, dose, reach, quality of implementation and the factors supporting implementation. Exploratory regression analyses will examine whether BEARS usage, contextual factors, implementation factors, and parent-reported child behaviour are associated with the primary outcome. Coding frameworks informed by the Technology Adoption Model [78] and the Technology Integration Model [79] will guide the deductive analysis of participant and parent/legal representative interviews alongside inductive coding to capture emergent data-driven themes. Deductive coding of clinician interviews will be informed by Normalisation Process Theory, alongside inductive coding to capture emergent data-driven themes. Data will be analyzed using Framework Analysis [76]. Following independent quantitative and qualitative analyses, findings will be integrated using the Triangulation Design–Convergence Model [80] to examine how implementation, mechanisms of impact, and contextual factors interact to influence intervention outcomes.

### Data monitoring and Ethics committee

The Data Monitoring and Ethics Committee (DMEC) is responsible for safeguarding the interests of trial participants, as well as monitoring the accumulating data and making recommendations to the Programme Steering Committee (PSC) on whether the trial should continue as planned. The DMEC will consider data in accordance with the statistical analysis plan and will advise the PSC through its Chair. There will be no interim analyses. Monitoring of the safety of the trial will be undertaken by the DMEC which will have untrammelled access to the trial data, and whose work is governed by a separate charter.

### Trial monitoring

The Clinical Trials Unit staff will review data and other information provided by investigators to identify trends, outliers, anomalies, protocol deviations and inconsistencies. Clinical trials unit staff will review CRF data for errors and missing key data points. The trial database will also be programmed to generate reports on errors and error rates. Essential trial issues, events and outputs, including defined key data points, will be detailed in the BEARS Data Management Plan.

### Protocol amendments

Protocol amendments are sent to each site’s research and development (R&D) department, PI and site coordinator and any other key members of staff on the delegation log via email with a description of the change and how the site will be affected. These are also communicated in sitewide bimonthly meetings and also the yearly investigators’ meeting. There have currently been seven substantial and fifteen non-substantial amendments approved by the REC/HRA for the BEARS trial [S3 File].

### Dissemination policy

A communications plan is in place to communicate all stages of this trial to all the various stakeholders, including patients and their families, CI centers, health service managers, educators, policymakers, and researchers etc. The communications plan aims to achieve and maintain recruitment, avoid resource waste by preventing research duplication, provide feedback to patients and the public about the use of public money, share new knowledge and learning generated, and contribute to potential implementation (scale-up) of the BEARS intervention after the project. We aim for targeted purposeful communications using the following channels: the BEARS website [81], social media, videos, promotional materials (e.g., badges, teddy key rings, etc.), press releases, conference presentations (including joint with patients), peer-reviewed and newsletter articles, plain language summaries, and public engagement events. The project has been built on PPIE, and this continues in our communications work. All communications acknowledge the NIHR, and we aim for all publications and associated data to be available in open access format.

The trial results will be disseminated regardless of the direction of effect. We also want to celebrate our achievements in:

- Engaging clinicians in a clinical trial (for the first time for many)
- Rolling out the BEARS speech perception tests to other clinics
- demonstrating innovative ideas for CI rehabilitation
- Involving deaf children and young people in research right from the start
- Using longitudinal qualitative methods to shape research design
- Empowering patients to take charge of their own rehabilitation
- Supporting any-time any-place CI rehabilitation with home tools

## Discussion

This RCT protocol has been designed to analyze whether using the spatial-listening training delivered via the BEARS training package for 3-months alongside usual care compared to only receiving usual care improves speech-in-noise perception, hearing experiences, vocabulary and quality of life in young people with bilateral CIs.

The underlying premise is that the BEARS training package will facilitate improvements in hearing abilities and listening skills, particularly in background noise, which in turn could improve self-confidence for engaging in communication without the fear of embarrassment. It is expected that vocabulary will be expanded, there will be improved concentration and attention, reduced anxiety and improved quality of life. Additionally, the measures used in the trial are intended to assess the direct and longer-term health and well-being outcomes as well as unpicking the mechanism of change.

The immersive VR format allows for home-based training, empowering children and young people with CIs to take more ownership of their rehabilitation and reducing the reliance on outpatient-based services. Moreover, the multi-modal format of the BEARS VR training package, which incorporates both audio and visual components, may be better than auditory interventions alone for generalization and retention [82].

The embedded health economic analysis in this RCT will provide evidence regarding the value for money of the BEARS compared to usual care alone in terms of health and quality-of-life benefits to CI users and the potential impact of this on their use of health care services. Importantly, it also incorporates the impact that these improvements in hearing and participation could have on local education support used by children and young people in the study.

Alongside the RCT, in-depth qualitative interviews and online, open-text surveys will capture the perspectives of children and young people regarding their experiences of listening in daily life and the impact of the BEARS training package on their real-life listening experiences. Qualitative findings will also contribute to our understanding of how the BEARS training package may lead to perceived changes in everyday listening for users of bilateral CIs. Focus groups with clinicians at the research sites conducted as part of the Process Evaluation will inform implementation strategies, and more generally, insights gathered from our qualitative research may inform new policy interventions, changes to clinical practice, and highlight opportunities to remove barriers to access and help promote well-being among children and young people with bilateral CIs.

Additionally, the trial’s use of iPad-based spatial speech-in-noise assessments highlights an opportunity to scale more efficient and accessible methods for spatial hearing assessment in routine clinical practice. Spatial speech-in-noise assessments (e.g., speech tests presented in noise where speech and noise come from different locations) provide a more realistic picture of how bilateral CI users function in noisy, real-world sound environments compared with standard pure tone audiometry [61]. However, these assessments are time consuming and have historically required expensive, maintenance-intensive multi-speaker arrays that impose significant space and operational costs. This has resulted in variable use of speech testing across the UK [83], which raises concerns about equality of access and uniformity of care. Delivery of the iPad-based speech tests in the BEARS trial demonstrates the possibility to use simple equipment to assess complex listening skills. Refining and translating these tools for clinical practice presents an opportunity to increase access to speech testing across geographical locations.

This is the first NIHR-funded multi-center hearing research study and is the largest known randomized controlled trial in children and young people with CIs. We are confident that the bespoke measures and the findings in this study will not only contribute to future audiological clinical practice and interventions but will also enable identification of further existing research gaps.

## Data Availability

No datasets were generated or analysed during the current study. All relevant data from this study will be made available upon study completion.

## Authors Contributions

[DV, DJ, NF, SD, LP, MM, CSC, NV-Z, FE, PK, HC] conceived of the study and developed the protocol, with input from [MS-C, BP, CR]. [NF, KC, NA, EB, CSC, DV, DJ] developed the statistical and health economic analyses plan. [DV, MM] wrote the first draft of the manuscript and subsequent revisions, with critical feedback from [LB]. [LA, JB, TH, EB, FK, FC, RN, SS] contributed to the refinement and finalisation of the study protocol and trial implementation. All authors contributed to the preparation of this manuscript and have reviewed and approved the final version.

## Acknowledgments

This study is funded by the NIHR Programme Grants for Applied Research Programme (NIHR201608). The views expressed are those of the author(s) and not necessarily those of the NIHR or the Department of Health and Social Care.

## Supporting Information

- **S1 File. SPIRIT Checklist**
- **S2 File. Study Protocol**
- **S3 File. Summary of Protocol Amendments**
- **S4 File. Pregnancy Notification Form**
- **S5 File. Statistical and health economics analysis plan**

